# Discrimination of Primary and Chronic CMV Infection based on Humoral Immune Profiles in Pregnancy

**DOI:** 10.1101/2024.02.21.24303056

**Authors:** Andrew P. Hederman, Christopher J. Remmel, Shilpee Sharma, Harini Natarajan, Joshua A. Weiner, Daniel Wrapp, Catherine Donner, Marie-Luce Delforge, Piera d’Angelo, Milena Furione, Chiara Fornara, Jason S. McLellan, Daniele Lilleri, Arnaud Marchant, Margaret E. Ackerman

## Abstract

Most humans have been infected by Cytomegalovirus (CMV) by the time they reach forty years of age. Whereas most of these CMV infections are well controlled by the immune system, congenital infection can lead to serious health effects and death for the fetus and neonate. With clear evidence that risk to the fetus is lower during chronic maternal infection, and varies in association with gestational age at the time of primary maternal infection, further research on humoral immune responses to primary CMV infection during pregnancy is needed. Here, systems serology tools were applied to characterize antibody responses to CMV infection among pregnant and non-pregnant women experiencing either primary or chronic infection. Whereas strikingly different antibody profiles were observed depending on infection status, more limited differences were associated with pregnancy status. Beyond known differences in IgM responses that are used clinically for identification of primary infection, distinctions observed in IgA and FcγR binding antibody responses and among viral antigen specificities accurately predicted infection status in a cross-sectional cohort. Leveraging machine learning, longitudinal samples were also used to define an immunological clock of CMV infection from primary to chronic states and predict time since primary infection with high accuracy. Humoral responses diverged over time in an antigen-specific manner, with IgG3 responses toward tegument decreasing over time as is typical of viral infections, while those directed to pentamer and glycoprotein B were lower during acute and greatest during chronic infection. In sum, this work provides new insights into the antibody response associated with CMV infection status in the context of pregnancy, revealing aspects of humoral immunity that have the potential to improve CMV diagnostics and to support clinical trials of interventions to reduce mother-to-fetus transmission of CMV.

## Introduction

A member of the herpesvirus family, human cytomegalovirus (CMV) commonly manifests as a mild or asymptomatic infection. However, in very young and immunocompromised individuals, CMV can cause severe disease; it is the leading cause of congenital infection among newborns^1^. In the United States alone it is estimated that 40,000 children are born with congenital CMV (cCMV) infection every year^1^. This high burden is still likely an underestimate since many cases are asymptomatic at birth. Many of the diagnosed infections are severe, leading to an estimated 400 deaths and an additional 8,000 cases presenting with permanent disabilities, including speech and language impairment, hearing loss, mental disability, cerebral palsy, and vision impairment, annually^1,2^. Fetal infection results from intrauterine transmission and is most likely to occur when a mother experiences primary CMV infection during pregnancy^3,4^. The difference in fetal infection risk between primary and reactivated maternal CMV infection is striking, with approximately one third of primary infections leading to CMV infection of the fetus as compared to < 3.5% estimated to result from CMV reactivation or superinfection^5-9^. New insights into CMV infection during pregnancy that could contribute to identification of pregnancies at greatest risk, and efficient testing of new therapeutic interventions are urgently needed^10^.

Pregnant women present a population of subjects who are not immunodeficient, but in a unique state of immune regulation in order to ensure fetal tolerance, and can transmit CMV to their fetus during pregnancy. While the differing risk of transmission associated with maternal seropositivity provides strong evidence that pre-conception immunity plays a protective role, studies evaluating the clinical potential of CMV-hyperimmune globulin to improve neonatal outcomes have yielded mixed results^11-13^. It has been speculated that these differing outcomes may relate to gestational age and the timing of the intervention following maternal infection and therefore be impacted by both the reliability of the diagnostic approach and strictness in the definition of primary infection cases. Because the suitability of these approaches for application during pregnancy can be supported by comparison of responses in pregnant with non-pregnant individuals during primary infection, developing a deeper understanding of humoral immune profiles in this unique immune state may have implications for the clinical development of diverse small molecule and biologic antiviral interventions.

Short of these goals, given the widely diverging congenital infection risks associated with primary and chronic infection, confident discrimination between these two states is crucial for identifying newborns with highest risk of cCMV infection. Complementing virological assessment, discrimination of these two states using serology may allow antiviral treatment in pregnant women and hearing and learning interventions in newborns to be initiated early^14,15^. Current diagnostic assays for pregnant women are performed using serology measurements for IgM and IgG avidity, while a PCR based urine test, often following a saliva PCR screening test, is the standard for newborns^14,16-18^. However, although CMV is a common infection, there is no universal standard diagnostic assay across countries and healthcare centers^19^, introducing variability in care and posing challenges to clinical trial design for testing of novel interventions. Additionally, many countries, including the United States, do not routinely screen for CMV in pregnant women, whereas some European countries do perform routine screening, adding another layer of complexity into clinical data regarding the impact of maternal CMV status, antibody responses, and timing of infection as assessed by differing diagnostic measures and across populations. Irrespective of these details, following a primary CMV diagnosis during pregnancy, clinicians typically counsel pregnant women about the future risks to their fetus.

The dichotomy in fetal risk profiles of primary and non-primary CMV infection has important implications in transmission risk for pregnant women. Beyond this classification, the specific timing of CMV infection is also a crucial factor in potential of severe congenital disease, with a primary infection in the first term of pregnancy leading to higher likelihood of severe cCMV infections as compared to a primary infection in the second or third trimester ^20^. Further insights into relative risk profiles are challenged by imperfect precision in defining and estimating the timing of primary infection, particularly during pregnancy, which has been associated with varying and dynamic immunological state changes^21^. As a result, more accurate and confident diagnosis of primary and non-primary CMV infection would benefit the medical community and affected birthing parents by providing clearer and more definitive insights into their CMV infection status and associated risk to the fetus.

However, previous work in a variety of infectious disease settings has shown that antibody responses evolve over time, exhibiting complex patterns in response magnitude and characteristics^22-24^, and that they can also differ in association with pregnancy^24-26^. Pregnant women with primary CMV infection represent a unique intersection of these two complex antibody response scenarios with established clinical significance. Here, leveraging machine learning and highly multiplexed assays to capture a wide range of antibody response attributes over time, we evaluate how responses to primary or non-primary CMV infection during pregnancy and over time may vary, providing insight into the role pregnancy may play in modifying these responses and new means of predicting and reducing risk of cCMV infection for affected pregnancies.

## Results

### Antibody Profiles Distinguish Primary from Chronic CMV Infection

Antibody responses were profiled among both pregnant and non-pregnant individuals with either primary or chronic CMV infection (**Table 1**), with primary infection strictly defined by CMV-specific IgG seroconversion, CMV-specific IgM antibody detection, low IgG avidity index, and/or CMV DNAemia. Conversely, chronic infection was defined by seropositivity in the absence of these diagnostic measures. To more comprehensively understand how antibody profiles vary among pregnant primary (n=114), non-pregnant primary (n=29), pregnant chronic (n=30), non-pregnant chronic (n=30) CMV-infected individuals, in addition to naïve subjects (n=9), antibodies specific for CMV tegument protein^27^ (CG1 and CG2), glycoprotein B (gB), and the pentamer complex were characterized for isotype, subclass, and Fc Receptor (FcR) binding capacity (**Supplemental Figure 1, Supplemental Table 1**). We profiled the antibody response examining a diverse set of antibody features and CMV antigens, including gB and pentamer complexes from several different sources, with the aim of gaining a more holistic view of the humoral immune response to primary or chronic CMV infection. Glycoproteins tested included several that are commercially available (Sino (Sino Biologicals), and NA (Native Antigen)), as well as others that have been characterized in structural studies conducted with an eye toward vaccine development (GSK)^28,29^, and by academic groups (UT)^30,31^, including a novel modified form of gB with engineered proline mutations (3p).

**Table 1.**
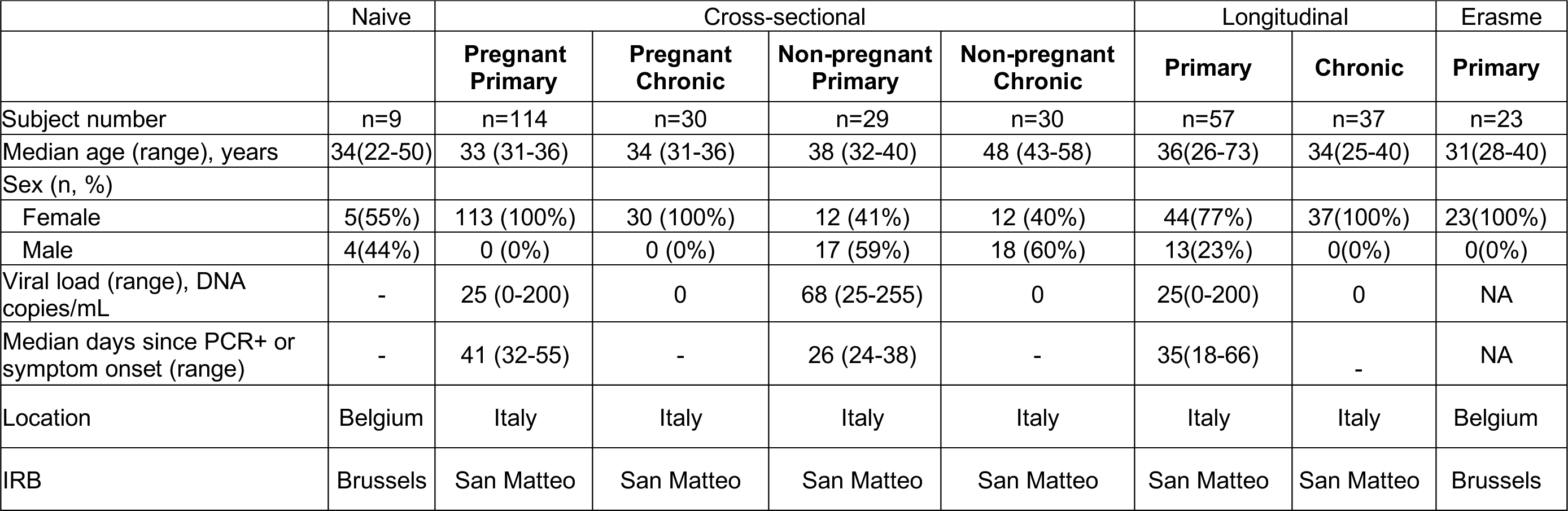
Cohort characteristics. Values that were not available are listed as NA.

To define differences in antibody profiles among subject groups that go beyond the measures typically used in clinical diagnosis, we performed UMAP analysis on CMV-specific antibody features excluding IgM **(Figure 1A)**. This unsupervised analysis revealed that the main aspects of differentiation among subjects related to infection status; strong distinctions in the antibody response were observed between individuals with primary as opposed to chronic infection. In contrast, limited differences in the global response profiles were observed between pregnant and non-pregnant individuals. While univariate analysis between pregnant and non-pregnant individuals did reveal some nominally statistically significant differences across or within primary and chronic infection groups (**Supplemental Figure 2**), we could not exclude the possibility that these differences resulted from a difference in timing of diagnosis or were impacted by differences in age between cohorts. The former may be likely, as similarly subtle differences were observed between primary subjects in these groups and another cohort (Erasme Hospital) of pregnant women with primary CMV infection (**Supplemental Figure 3**). Given these observations and the lack of robust differences associated with pregnancy status, this variable was not considered in subsequent analyses.

**Figure 1:**
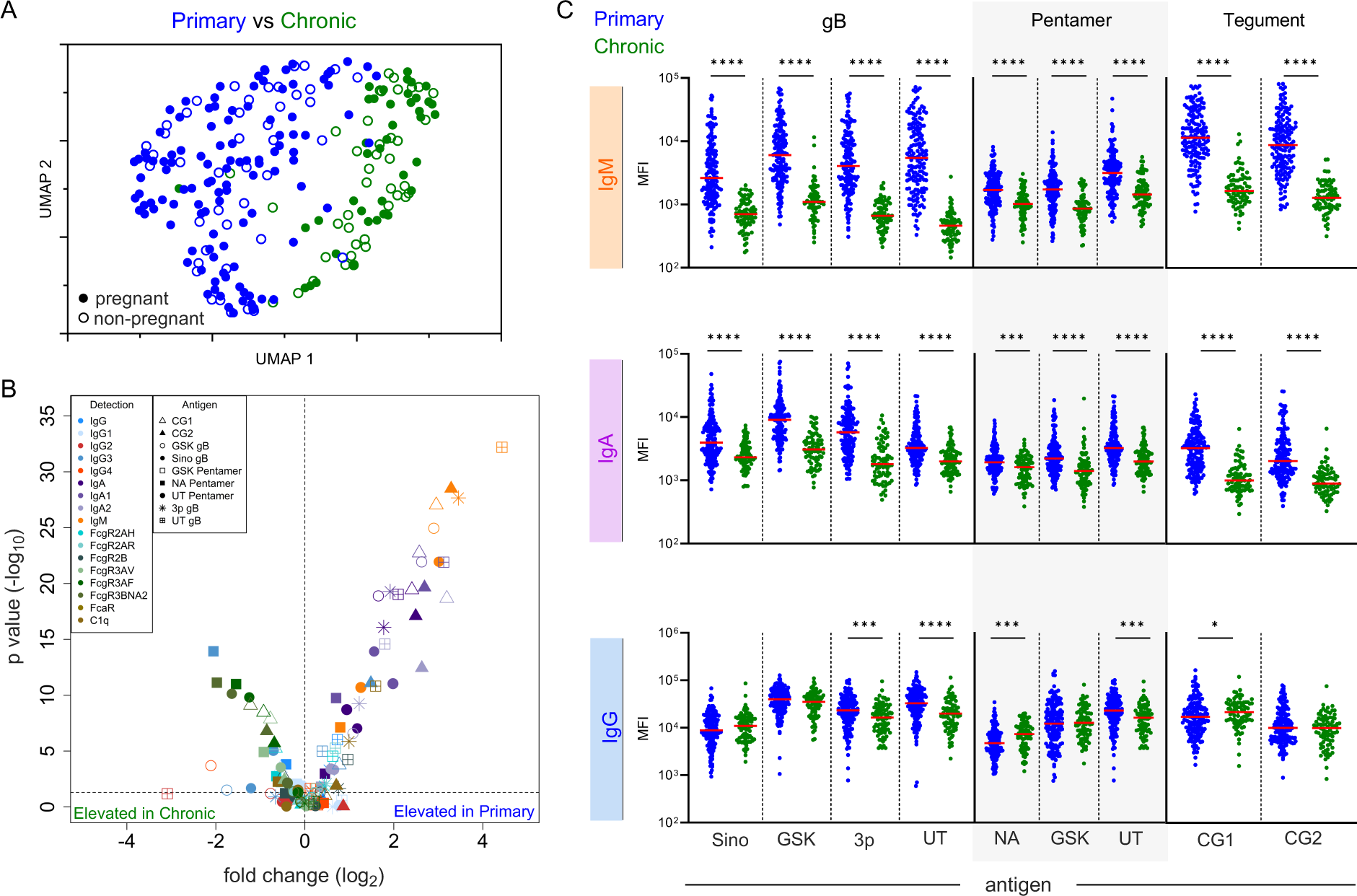
Antibody features distinguish primary from Chronic CMV infection but not pregnancy status. **A.** Uniform manifold approximation (UMAP) biplot of antibody features excluding IgM. Distinct clusters of subjects with primary (blue) and chronic (green) infection but not pregnancy status (hollow and filled symbols) are observed. **B.** Volcano plot of each CMV-specific antibody feature assessed. Volcano plot represents the log_2_ fold change (x-axis) against the –log_10_ p value (Mann Whitney test: *p < 0.05, **p < 0.01, ***p < 0.001, and ****p < 0.0001). Antibody specificities (Antigen) are indicated by shape and Fc characteristics (Detection) indicated by color. **C.** IgM, IgA, and IgG binding to CMV antigens (further described in Supplemental Table 1). Data are the mean median fluorescent intensity (MFI) values of technical replicates. Solid red line indicates median.

We next investigated which individual features of the immune response might contribute to clustering of subjects in primary and chronic infection groups. Visualizing the degree and confidence of differences in antibody response features between primary and chronic infection status groups revealed aspects of the humoral response that consistently differed (**Figure 1B-C)**. Distinct levels of CMV-specific IgA, IgM, and FcR binding antibodies were observed between primary and chronic infection groups across multiple CMV antigens. IgM responses were elevated across all antigens for the primary infection group, which was expected as IgM titers were used to define infection status. Interestingly, total CMV-specific IgA and both IgA1 and IgA2 subclasses were elevated in primary infection subjects. Somewhat surprisingly, total IgG had very modest differences between primary and chronic infection groups.

Given the striking differences among groups we next wanted to further examine individual responses for IgM, IgA, and IgG across the panel of CMV antigens. As expected, IgM responses were strongly and significantly elevated among subjects with primary infection, as were IgA responses across diverse antigen specificities (**Figure 1C**). Interestingly, and in contrast with total IgG response levels (**Figure 1C**), which exhibited statistically significant but relatively small differences in response magnitude, FcγR-binding antibodies were both significantly and strongly elevated among subjects with chronic CMV infection (**Figure 1B, Supplemental Figure 4**), suggesting the presence of qualitative differences in the antibodies present during primary and chronic infection.

### IgG Subclasses and CMV Antigens Display Distinct Profiles in Primary and Chronic Infection

The observation that FcγR binding antibodies but not total IgG were reliably elevated in chronic subjects was intriguing, and suggested that differences in induction of IgG subclasses, which differ dramatically in their FcR binding capacity, may exist. However, with the exception of IgG2 responses to tegument, IgG2 and IgG4 responses to CMV antigens were uncommon, and therefore differences in the levels of these more functionally inert subclasses were not observed (**Supplemental Figure 5**). Humoral responses to viral infections are generally dominated by IgG1 and IgG3 antibodies, which both bind well to activating FcγR and have the potential to elicit the potent antiviral activities of the complement cascade and innate immune effector cells. While statistically significant differences were observed for a subset of antigens tested, IgG1 responses, which are typically dominant following acute viral infections^32,33^, were generally similar in primary and chronic infection (**Figure 2A**). Only one of each of the gB, pentamer, and tegument proteins tested showed a statistically significant difference, and each of these demonstrated only a small increase among the chronic infection group. In contrast, IgG3 responses were more uniformly and strikingly distinct between groups. Perhaps most surprisingly, the direction of these differences varied by antigen-specificity. Antibody responses to tegument proteins exhibited elevated levels in primary infection, but elevated levels of gB and pentamer-specific IgG3 were observed among the chronic infection group (**Figure 2A**). While all pentamer complexes tested showed this profile, responses to recombinant gB proteins were more variable, with two gB preparations showing elevated levels among chronic subjects, and one preparation showing elevated levels in primary infection. While some distinctions in IgG subclass composition in the context of primary and latent herpesvirus infections have been reported^34-38^, how well they may support discrimination between primary and chronic CMV infection is not known. In sum, while IgG1 responses tended to either persist or increase between primary and latent infection classes, IgG3 responses differed dramatically by antigen-specificity; responses to tegument were higher among subjects with primary infection, but responses to gB and pentamer were instead elevated in chronic infection.

**Figure 2:**
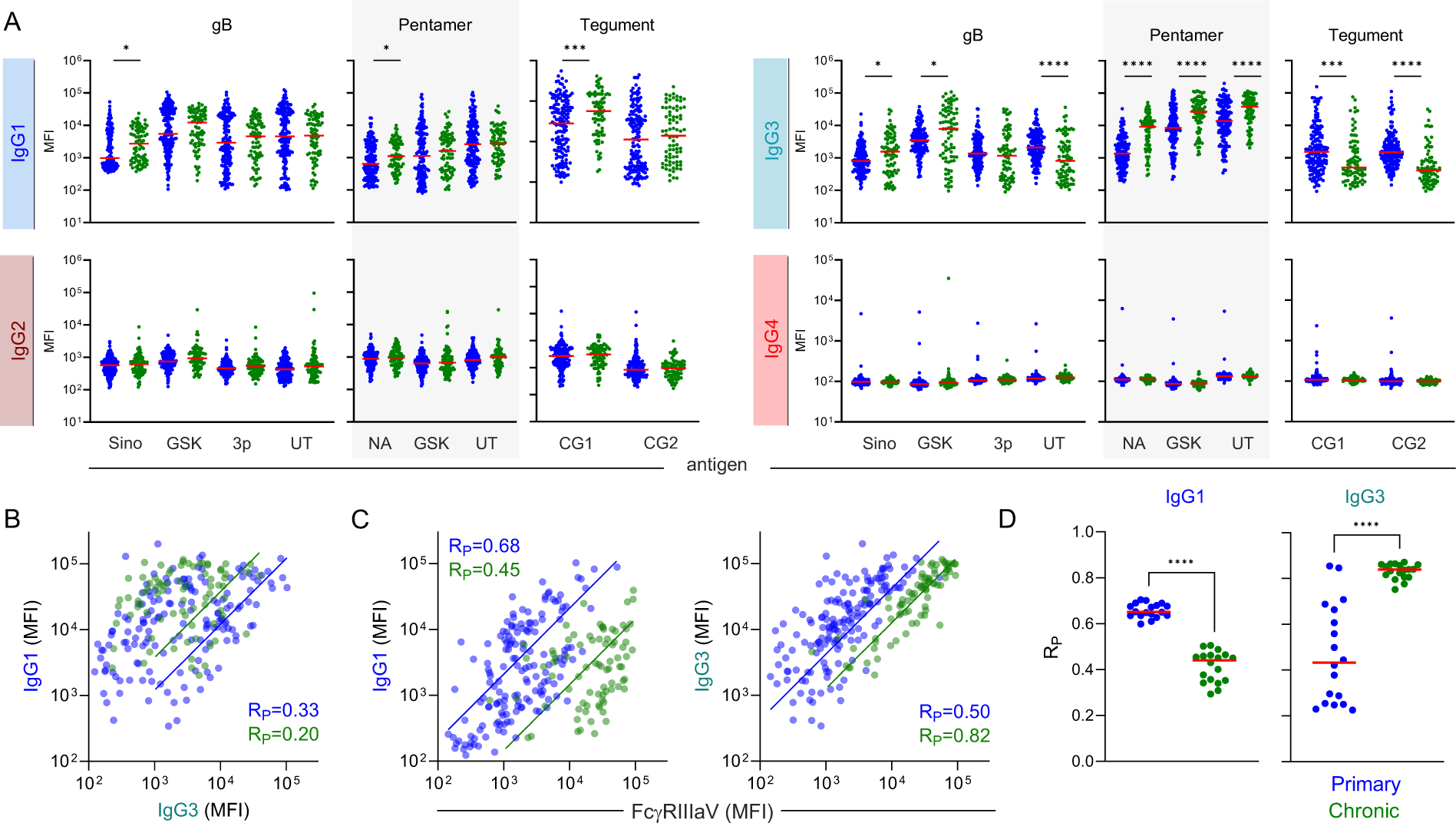
IgG Subclasses display distinct profiles in primary and chronic infection depending on antigen specificity. **A.** Levels of gB-, pentamer-, and tegument-specific IgG1, IgG2, IgG3, and IgG4 antibodies in individual with primary (blue) or chronic (green) CMV infection. **B.** Scatterplot and fit lines of levels of (UT) pentamer-specific IgG1 and IgG3 responses by infection status. Pearson correlation coefficient (R_P_) indicated in inset**. C.** Scatterplot and fit line of levels of (UT) pentamer-specific IgG1 (left) and IgG3 (right) versus (UT) pentamer-specific FcγRIIIaV-binding antibodies. Pearson correlation coefficient (R_P_) indicated in inset**. D.** Pearson correlation coefficients (R_P_) for each pentamer antigen tested across FcγR by infection status. Solid lines denote group medians; differences between groups were assessed by Mann-Whitney test (*p < 0.05, **p < 0.01, ***p < 0.001, and ****p <0.0001); values presented are median fluorescent intensities (MFI).

While UT pentamer-specific IgG1 and IgG3 levels were correlated with each other among subjects with primary infection (R_P_=0.33, p<0.0001), they were not well correlated among individuals with chronic infection (R_P_=0.20, p=0.08). This difference in degree of correlation between IgG1 and IgG3 responses between infection status states was consistent across pentamer proteins evaluated. Additionally, the strength of correlation observed between these IgG subclasses and the FcγRIIIaV-binding capacity of pentamer-specific antibodies changed over time, with stronger correlations between IgG1 and FcγRIIIaV observed in primary infection, but IgG3 and FcγRIIIaV binding in chronic infection (**Figure 2C**). These patterns were consistent across FcγRs and pentamer proteins tested (**Figure 2D**), suggesting that the pool of antibodies most capable of eliciting FcγR-dependent effector functions changes in composition over the course of infection. These temporal differences may have important implications to viral pathogenesis and host defense.

### Prediction of CMV infection status using machine learning

Next, because unsupervised analysis of antibody profiles revealed clear distinctions between primary and chronic CMV infection, we applied supervised machine learning to explore the ability of a model to accurately discriminate primary and chronic CMV infection (**Figure 3A**). We employed a logistic regression framework with regularization to classify primary or chronic CMV infection based on antibody profiling data while reducing the risk of overfitting associated with high-dimensional data. Again, IgM responses were excluded because they were used in clinical class assignment of the study groups. The model was trained on 80% of primary and chronic subject profiles while the remaining 20% was used for testing. Furthermore, repeated five-fold cross validation was employed so each subject would be part of the test set and representative accuracy across different folds could be defined.

**Figure 3:**
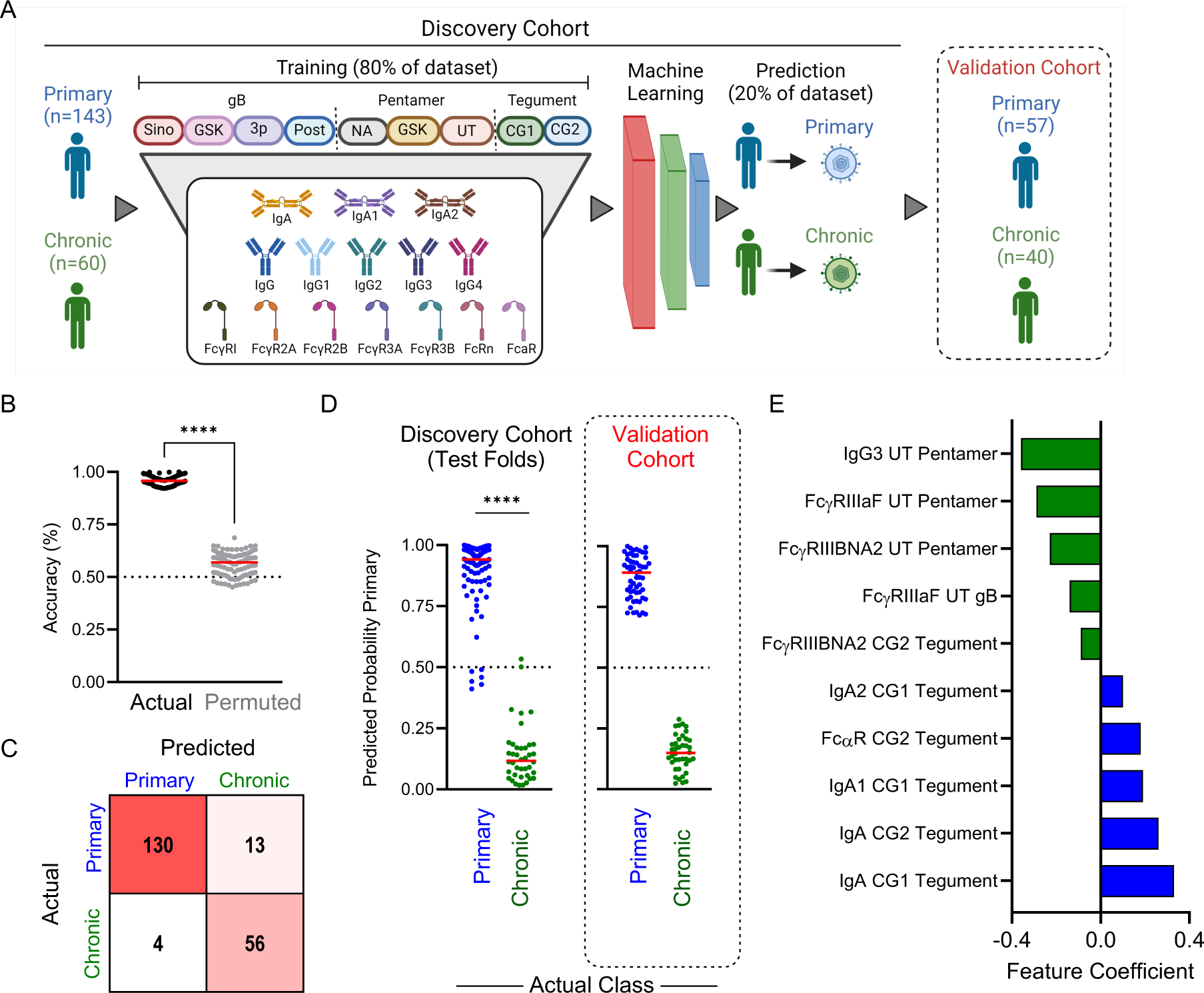
Machine learning accurately predicts primary or chronic CMV infection status. **A.** Schematic overview of cross-validated machine learning workflow employing antibody profiling data to discriminate between primary and chronic infection status in discovery and validation cohorts. **B.** Prediction accuracy for 100 repeated 5-fold cross-validation runs on actual (black) and permuted (gray) class labels. **C**. Confusion matrix of predicted versus actual class labels in the median model for 5-fold cross validation. **D.** Class probabilities of each sample in the discovery set when evaluated as a test sample in the cross-validation run exhibiting median performance (left) and for the validation cohort using the final model (right). **E.** The identities and coefficients of the features making the largest positive (n=5) or negative (n=5) contributions to the final model. Solid lines denote group medians; differences between groups or conditions were assessed by Mann-Whitney test (*p < 0.05, **p < 0.01, ***p < 0.001, and ****p <0.0001).

Model predictions were highly accurate; across 100 repeated five-fold cross validation runs, the median model accuracy for predicting primary or chronic CMV infection was 94% (**Figure 3B**). In contrast, prediction results were essentially random when training and testing was performed after permutation of class labels, which serves as a means to assess model robustness by measuring the potential for overfitting. Misclassifications were not biased toward one or the other class, as shown for the cross validation run repeat presenting median accuracy as a representative confusion matrix (**Figure 3C**). Classification calls for actual but not permuted class labels were both typically correct and assigned to respective classes with high probability (**Figure 3D**). The relatively few incorrect classifications were typically close to the decision boundary. Given this evidence of model accuracy and robustness, a final model was trained on all subjects in the discovery cohort. When applied to an independent set of 57 primary infection and 40 chronic infection samples serving as a validation cohort, this model resulted in similarly high confidence and perfect classification accuracy (**Figure 3D**). Given this validation, the top features employed in the final model were examined (**Figure 3E**). The features with largest positive coefficients, which serve to identify primary subjects, were primarily IgA-related antibody responses directed to tegument antigens (4/5 features). Conversely, features with large negative coefficients, useful to identify chronic infection, were typically related to IgG3 and FcγRIII-binding capacity of response to gB and pentamer (4/5 features). Collectively, these modeling results point to novel antibody response attributes, distinct from traditionally considered parameters such as IgM and IgG avidity, as being excellent candidate markers for distinguishing primary and chronic infection status.

### Modeling Longitudinal Responses to CMV Infection Reveals a Molecular Clock of Antibody Responses

The excellent discrimination of primary and chronic CMV infection in the validation cohort led us to next explore how class predictions related to longitudinal development of humoral immune responses to infection. To this end, subsequent samples were available for the validation cohort over a series of up to four visits that extended out to half a year after initial sampling or infection onset. The model trained on the initial cohort was used to predict class for longitudinal samples from the validation cohort (**Figure 4A**). Whereas chronic subjects were consistently classified as such with similarly high confidence at all subsequent visits, subjects with primary infection at the initial timepoint became less confidently classified as primary at subsequent visits (**Figure 4B**). By visit four, the majority of samples from subjects with primary infection at their initial visit exhibited class probabilities below the lowest observed at the initial sampling. However, because visits were not consistently spaced in time between subjects, these longitudinal profiles can be more meaningfully compared over time post symptom onset (**Figure 4C**). Despite the imperfect reliability of projected timing of infection, the classification model probabilities presented a clear relationship with time. Samples fell below the midpoint on the classification scale as early as 90 days post infection onset, but failed to reach values typically observed among chronic subjects even out at 250 days post infection, suggesting a relatively prolonged transition to reaching the chronic state profile. As expected, the individual antibody response features making the greatest contributions to the classification model also demonstrated clear changes among subjects with primary, but not chronic infection, over time (**Figure 4D**).

**Figure 4:**
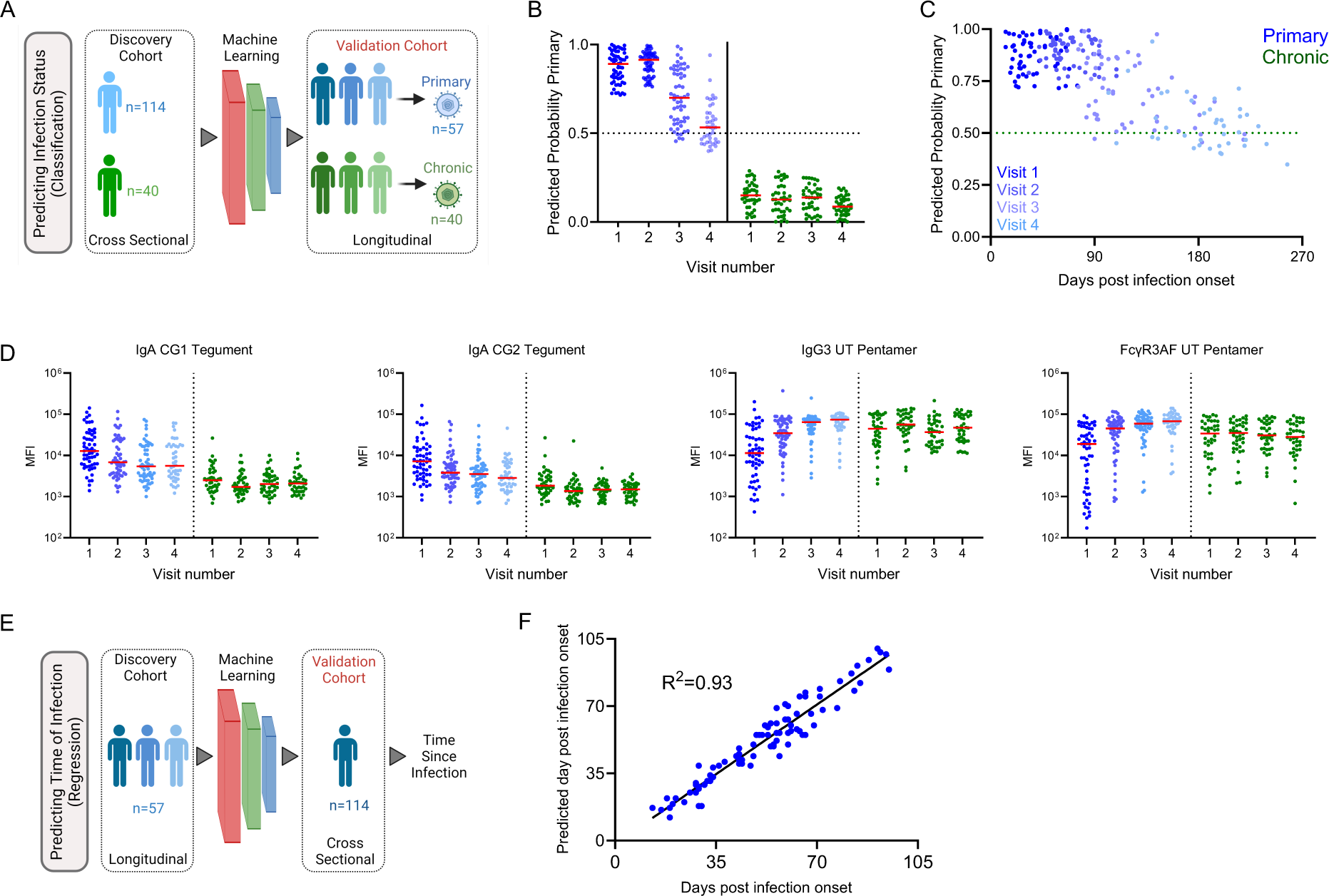
Longitudinal models define a molecular clock of CMV primary infection. **A.** Analysis overview. The infection status classification model trained on the cross-sectional cohort was applied to longitudinal samples available from the validation cohort. **B.** Class probabilities of each sample in longitudinal cohort over sample collection visits for individuals with primary (blue) and chronic (green) infection. **C**. Scatterplot of class probabilities for subjects defined as having primary infection at visit 1 over time. **D.** Scatterplots of features employed by classification model to predict infection status over time in the longitudinal cohort. **E**. Analysis overview. Primary infection samples from the longitudinal cohort samples were used to train a regression model to predict time since infection (days post symptom onset) that was applied to the primary samples from the cross-sectional cohort. **F**. Scatterplot of model predictions of time since infection when primary samples used for predicting days post symptom onset. Data shows the measure of the predicted label and its closeness to the true label.

To this point, machine learning models have only been concerned with making predictions on the probability of a sample belonging to either the primary or chronic infection class. However, given the clear ability for these binary classification models to provide insight into time since infection, we next sought to evaluate models explicitly trained for this specific purpose. For this purpose, longitudinal profiles of the primary infection cases across visits were used to train a model to predict time since symptom onset as a continuous variable (**Figure 4E**). The resulting linear regression model, which showed good robustness in the context of five-fold cross-validation (**Supplemental Figure 6A**), was then used to predict days post infection for the cross-sectional primary samples. This model, which relied primarily on IgG3 features (**Supplemental Figure 6B**) showing strong time-dependence (**Supplemental Figure 6C**), was then applied to the cross-sectional cohort. Predictions of time since symptom onset in the unseen validation cohort exhibited excellent accuracy (**Figure 4F**). Overall, this analysis demonstrated that antibody profiles in CMV-infected individuals exhibit generalizable temporal patterns in their dynamic antibody responses during primary infection.

## Discussion

Presently, discrimination of primary and non-primary CMV infection in the context of pregnancy is used in counseling as to the risk of congenital CMV infection based on the lower risk of congenital infection associated with non-primary infection and the prescription of antiviral therapies. Beyond this value, in the absence of an effective vaccine, a deeper understanding of how immune responses differ in association with transmission risk has the potential to contribute to the development of new interventions. Here, high dimensional antibody profiles beyond IgM levels and IgG avidity were developed from a commonly used multiplex assay format and supervised and unsupervised machine learning was used to differentiate primary and chronic infection and pregnancy status.

Whereas limited or no differences in humoral responses were associated with pregnancy status, our study showed clear distinctions in antibody profiles between primary and chronic infection cases. Both composite profiles individual antibody features related to antigen-specificity and immunoglobulin isotype, subclass, and binding to Fc receptors demonstrated these distinctions. Importantly, these differences extended beyond those previously known to exist and which are presently applied to support clinical diagnosis. The number and longitudinal profiles of the features that distinguish infection history suggested that a sort of humoral clock could be defined in order to time the onset of primary CMV infection. Indeed, supervised machine learning models that reliably captured how antibody responses to CMV infection varied over time were learned and validated on independent samples.

Beyond potential clinical utility, the defining features of how responses to primary CMV infection transition over time is relevant to understanding the evolution of the immune response to this member of the notoriously immune-evasive Herpesviridae family^39,40^. Current diagnostic methods rely on detection/levels of IgM and of IgG avidity. The presence of IgM alone is insufficient to diagnose a primary CMV infection: poor correlation between commercial tests have been reported^41^, and false positive primary status calls can result from both persistence of IgM as well as boosting in response to reactivation^42,43^. Likewise, a positive test for CMV-specific IgG indicates an individual is positive for CMV but provides little information into the time since infection^14,44^. However, the combination of IgM positivity and low IgG avidity is generally considered to be a reliable indicator of primary CMV infection, though interpretation of IgG avidity tests are confounded by low levels of CMV-specific IgG, intermediate responses raise classification issues, and these indicators are supported by clinical studies of small size. Coupled to the lack of an international standard serum panel of samples from primary infection, the difficulty inherent to establishing a uniform, accurate, and robust diagnostic method is clear^45^. Our data demonstrates that there are other changes in the secreted antibody repertoire that reliably occur over consistent time periods during primary infection.

Prior studies of IgG subclasses of CMV-specific antibodies have consistently reported the IgG1 and IgG3 bias typical of anti-viral antibodies but noted that subclass responses and total IgG titers can be discordant^35,46^. Further, they have attributed relatively greater neutralization potency to this minor portion to IgG3^36^ To that end, we were surprised to observe increasing levels of IgG3 specific for viral glycoproteins over time. Whereas IgG3 responses are usually associated with acute infections and typically wane over time^33,47-49^, they were observed to increase across both cross-sectional and longitudinal cohorts, but to do so in an antigen-specific manner. Namely, whereas IgG3 responses to tegument proteins showed the typical pattern and decreased while IgG1 responses were stable or increased, IgG3 responses to pentamer increased while IgG1 responses remained largely stable. Memory B cells (MBC) specific for gB are known to exhibit phenotypic states distinct from those specific for tegument antigens^50^. The reduced frequency of MBCs with effector potential specific for glycoprotein as compared to tegument observed in this prior study may relate to the altered kinetics of IgG subclasses observed here. While the mechanistic underpinning and the biological implications of this unusual pattern have yet to be determined, CMV expresses two Fc binding proteins, gp34 and gp68^51^. Unlike the viral FcR of HSV, these proteins have been reported to bind to all human IgG subclasses^52,53^, though affinities have not been reported. Additionally, how these two surface proteins contribute to viral pathogenesis is incompletely understood^54^.

Limitations of this study include the fact that the majority of samples were sourced from a single geographic region, leaving open the possibility that differences associated with host genetics and environmental and infectious disease exposure history impact these observations. While relationships between maternal immune responses and transmission of cCMV are of exceptionally high clinical relevance, this study focused on CMV infection history. However, while risk is considerably greater during primary infection, there are certainly differences in virologic, innate immune, and other factors that contribute to these differing risk profiles^55-58^. The influence of maternal antibody responses is unclear, particularly given the conflicting results in studies of passive antibody therapy in the context of primary maternal infection^59,60^. While the dosing and frequency of hyperimmune globulin also differed in these studies, a positive effect was only observed in women with very early infection, pointing to the potential importance of accuracy in the dating of infection recency. Recent studies have started to explore the role that antibody responses play in CMV transmission in a rhesus macaque model^61,62^. However, data in humans is limited and often confounded by the differing risk of infection associated with primary as compared to non-primary infection^63^. Additionally, the panel of antigens tested in our investigation was not exhaustive, and the reactivity patterns sometimes differed in association with different sequences, conformational states, expression hosts, and other factors, the contributions and importance of which have yet to be defined. These and other factors may represent worthwhile directions for a future study.

Overall, many open questions regarding the role of humoral immunity in the context of CMV infection, transmission, latency, and reactivation remain. Higher resolution and more comprehensive analysis of antibody responses using systems serology approaches has the potential to improve our understanding of the complex virus-host interactions at play. Here, by analyzing highly curated cohorts, we report and validate phenotypic signatures of gB, pentamer, and tegument-specific antibody responses that not only robustly classify primary infection status, but also provide insights into time of infection. It remains to be seen if the atypical dynamic profile of IgG3 responses to envelope glycoproteins elicited by CMV is antigen-intrinsic and might be recapitulated when these antigens are delivered by other means, or if it may represent an evasion strategy dependent on other viral genes or aspects of the innate response to viral infection. In the meantime, this work stands to define novel hallmarks of primary CMV infection and time of infection that may present new opportunities to streamline primary infection diagnosis, potentially impacting current clinical practice and enrolment of pregnant women with primary infection in interventional trials, thereby providing new insights into relative cCMV risk and management strategies.

## Methods

### Clinical Samples

Serum samples were gathered from subject cohort groups of pregnant primary infection, pregnant latent infection, non-pregnant primary infection, non-pregnant latent infection, as well as CMV negative patient cohort as a negative control group (**Table 1**). Human subjects were recruited from Fondazione IRCCS Policlinico San Matteo, Pavia, Italy, and included healthy, primary, and latent CMV infected subjects, as well as pregnant and non-pregnant subjects. An additional cohort of pregnant women with primary infection was recruited from Erasme Hospital. Diagnosis of primary HCMV infection was based on two or more of the following criteria: CMV-specific IgG seroconversion, CMV-specific IgM antibody detection, low IgG avidity index and CMV DNAemia. Chronic infection was defined by the presence of CMV-specific IgG and absence of CMV-specific IgM antibody, and no detection of CMV DNA in blood, saliva, urine and genital secretion during pregnancy. In 89/114 (78%) pregnant women and 28/29 (95%) non-pregnant subjects, onset of primary infection was defined by the appearance of symptoms, while in 19/114 (17%) asymptomatic pregnant women onset of infection was estimated on seroconversion (i.e. in the mid between the last IgG negative and the first IgG positive result), occurring within a ≤6-week interval. Finally, in six asymptomatic pregnant women (5%) and one (3%) non-pregnant subject, onset of infection was estimated on the kinetics of CMV-specific IgM and IgG avidity index. For a subgroup of 40 pregnant women and 28 non-pregnant subjects with primary infection, 2-4 sequential serum samples collected until 6 months after onset of infection were available. Samples from pregnant women with chronic infection were collected at 10, 20, and 30 weeks of gestation and at delivery. Sample collection was approved by Fondazione IRCCS Policlinico San Matteo, and each participant gave written informed consent.

### Antibody Profiling Experiments

Fc receptors were expressed as previously described^64^. A complete list of Fc detection reagents and antigens used is provided in **Supplemental Table 1**. Pentamer and glycoprotein B were sourced both commercially, through industry partners, and expressed in-house. For the latter, HCMV Pentamer (UT, Towne strain) was produced by mixing plasmids encoding for residues 24 to 718 of gH with a C-terminal 6× HisTag, residues 31 to 278 of gL, residues 21 to 171 of UL128, residues 26 to 214 of UL130, and residues 19 to 129 of UL131A, all with artificial signal sequences, at an equimolar ratio^31^. This mixture was then used to transiently transfect FreeStyle293-F cells via polyethylenimine. Plasmids encoding for postfusion HCMV gB (AD169 strain) residues 32–692 with an artificial N-terminal signal sequence and a C-terminal HRV3C protease cleavage site, 8×HisTag, and a TwinStrep tag. One postfusion gB construct (UT, or JSM-1074) contained substitutions Y155G, I156H, Y157R, Y206H, S238N, W240T, L241T, Y242H and C246S, which have been reported previously^30^ and the other construct (3p, or JSM-956) contained substitutions Y155G, I156H, Y157R, Y206H, W240A, L241T, Y242H, C246S, R456A, R459G, M472P, R491P, G492P and a C-terminal T4 fibritin trimerization motif. Plasmids were transiently transfected into FreeStyle 293-F cells via polyethylenimine. Pentamer was purified using NiNTA resin (Thermo Scientific) and postfusion gB proteins were purified using Strep-Tactin affinity resin (IBA Lifesciences). Affinity-purified proteins were further purified by size-exclusion chromatography using a Superose6 10/300 column (Cytiva) in a buffer composed of 2 mM Tris pH 8.0, 200 mM NaCl, 0.02% NaN3.

Characterization of antibody profiles was performed using the Fc array assay^65,66^. Antigens were covalently coupled to magnetic microspheres (Luminex Corporation) using carbodiimide chemistry. Serum dilutions used in assays ranged from 1:250-1:5000 based on initial pilot experiments and previous experience. Detection of antigen specific antibodies was done using R-phycoerythrin-conjugated secondary reagents specific to human immunoglobulin isotypes and subclasses and by Fc receptor tetramers. Median fluorescent intensity data was acquired on a FlexMap 3D array reader (Luminex Corporation). Samples were run in technical duplicates and results were averaged.

### Classification of CMV infection status

A binomial logistic regression model with least absolute shrinkage and selection operator (LASSO) regularization was used to prediction infection status. Model training was performed using the scikit-learn (version 1.3) in Python (version 3.9) with default options. The regularization parameter was chosen using the option that gave the lowest classification error. Model accuracy was determined by the test set label predictions compared with true labels. Accuracy was assessed over 100 repetitions of five-fold cross validation. Permutation testing was done to measure model robustness by doing the same procedure as described but on randomly shuffled data. Feature importance was determined from a final model that included all subjects.

### Prediction of time since infection

The same machine learning model as described for predicting CMV infection status was used, this time minimizing mean squared error in time since infection. The model was trained on cross sectional data and tested on all longitudinal samples.

## Statistics

Statistical analysis was performed in GraphPad Prism (version 9.7). UMAP plots were generated in Python (version 3.9) using the umap-learn package (version 0.4)^67^ and then plotted using Prism. Volcano plots were generated in R (version 4.3) using ggplot2. Statistical tests are described in the relevant figure legends.

## Study approval

The study was approved by the IRBs at Fondazione IRCCS Policlinico San Matteo and Erasme Hospital for sample collection, and Dartmouth College for sample testing and analysis.

## Data availability

Data used in the study is available as **Supplemental Data File 1**.

## Data Availability

Data used in the study will be made available as Supplemental Data File 1 upon publication in peer-reviewed forum.

## Acknowledgements

This study was supported in part by Infect-ERA, GSK and Sanofi via the CYMAF consortium, as well as by NIH U19AI45825. Pentamer and gB proteins were provided by GSK. A.M. is Research Director at the F.R.S., FNRS, Belgium.

