## Supplemental Figures and Tables for "Discrimination of Primary and Chronic CMV Infection based on Humoral Immune Profiles in Pregnancy"

**Supplementary Materials**

| **Supplementary Figures** | |
|---|---|
| Supplementary Figure 1 | Heatmap of antibody responses. |
| Supplementary Figure 2 | Comparison of antibody responses in pregnant and non-pregnant individuals. |
| Supplementary Figure 3 | Comparison of antibody responses in pregnant individuals from Erasme and those from other medical centers. |
| Supplementary Figure 4 | Representative boxplots of CMV-specific antibody binding to FcγR across groups. |
| Supplementary Figure 5 | Representative boxplots of CMV-specific IgG subclasses across groups. |
| Supplementary Figure 6 | Longitudinal machine learning model performance and feature importance |
| **Supplementary Tables** | |
| Supplementary Table 1 | Fc detection and antigen reagents. |

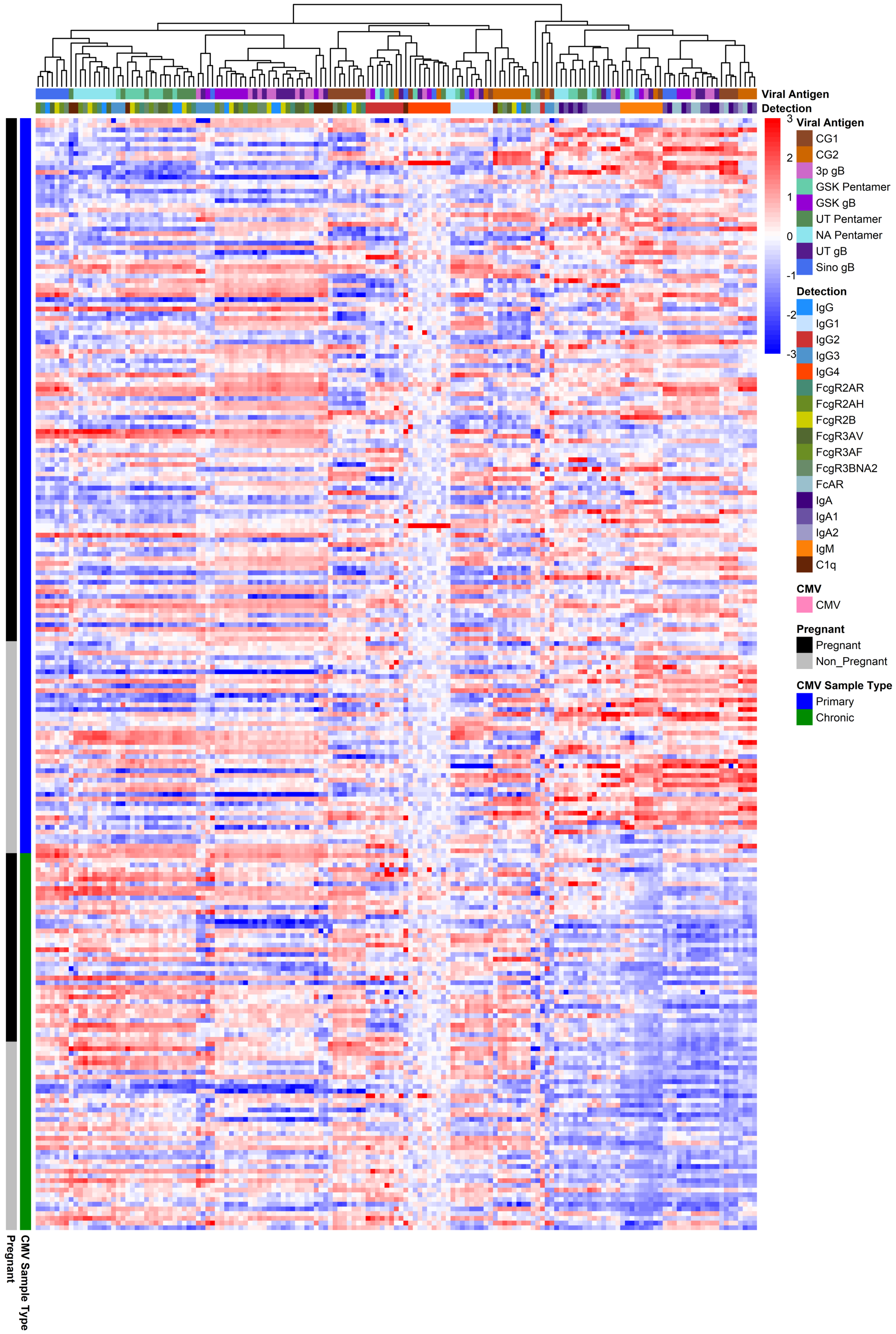


**Supplemental Figure 1: Heatmap of hierarchically clustered CMV-specific Fc array features across subjects in the cross-sectional cohort**. Each row represents an individual subject. Subjects are grouped by status, as indicated by the vertical color bars. Each column represents an Fc array feature; horizontal color bars indicate each function or each Fv-specificity (Antigen) and Fc-characteristic (Detection) tested. Responses are scaled and centered per feature and the range was truncated ± 3 SD.

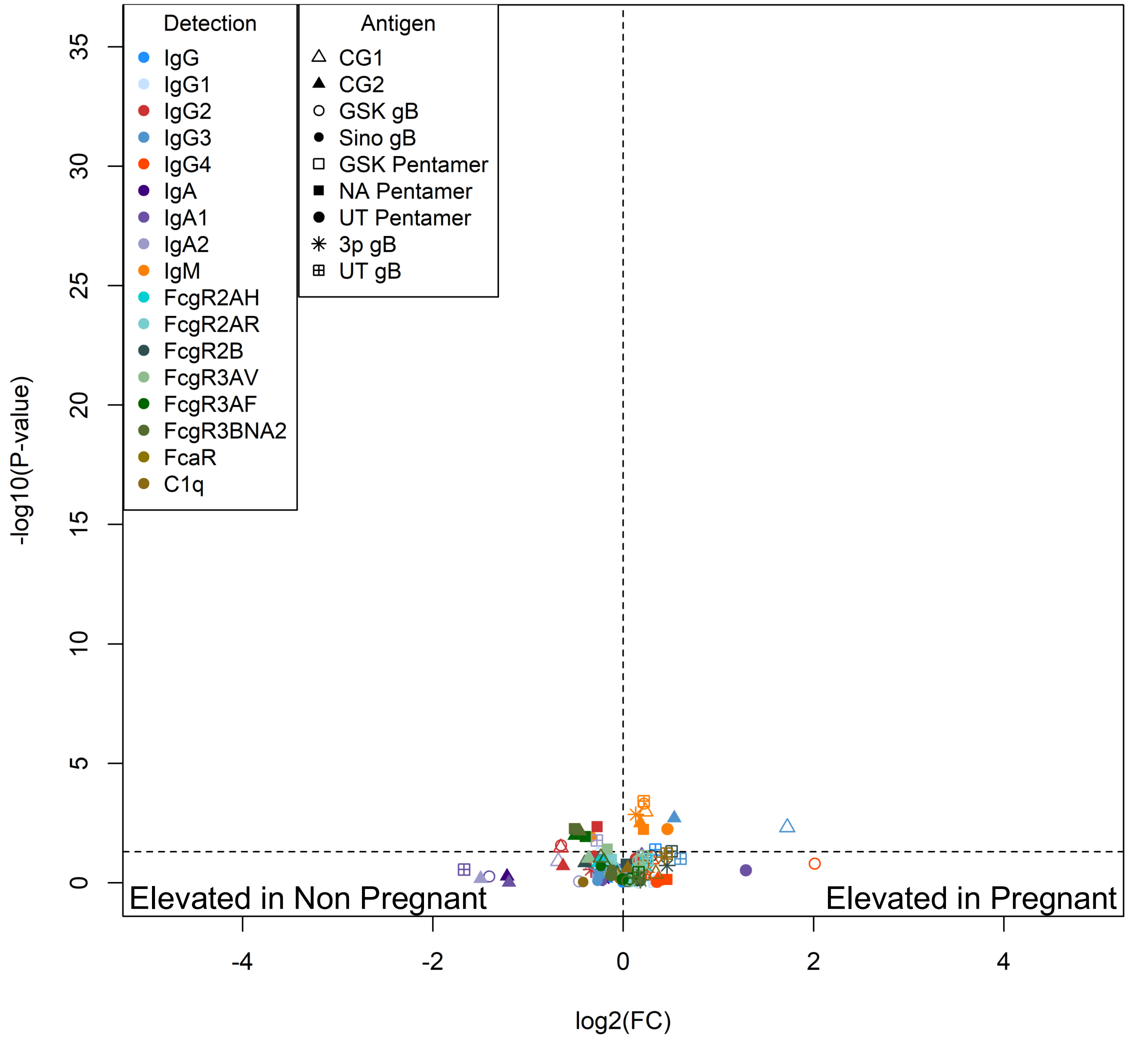


**Supplemental Figure 2: Comparison of pregnant and non-pregnant antibody responses**
Volcano plot of each CMV-specific antibody feature assessed. Volcano plot represents the log2 fold change (x-axis) against the –log10 p value (Mann Whitney test). Antibody specificities (Antigen) are indicated by shape and Fc characteristics (Detection) indicated by color.

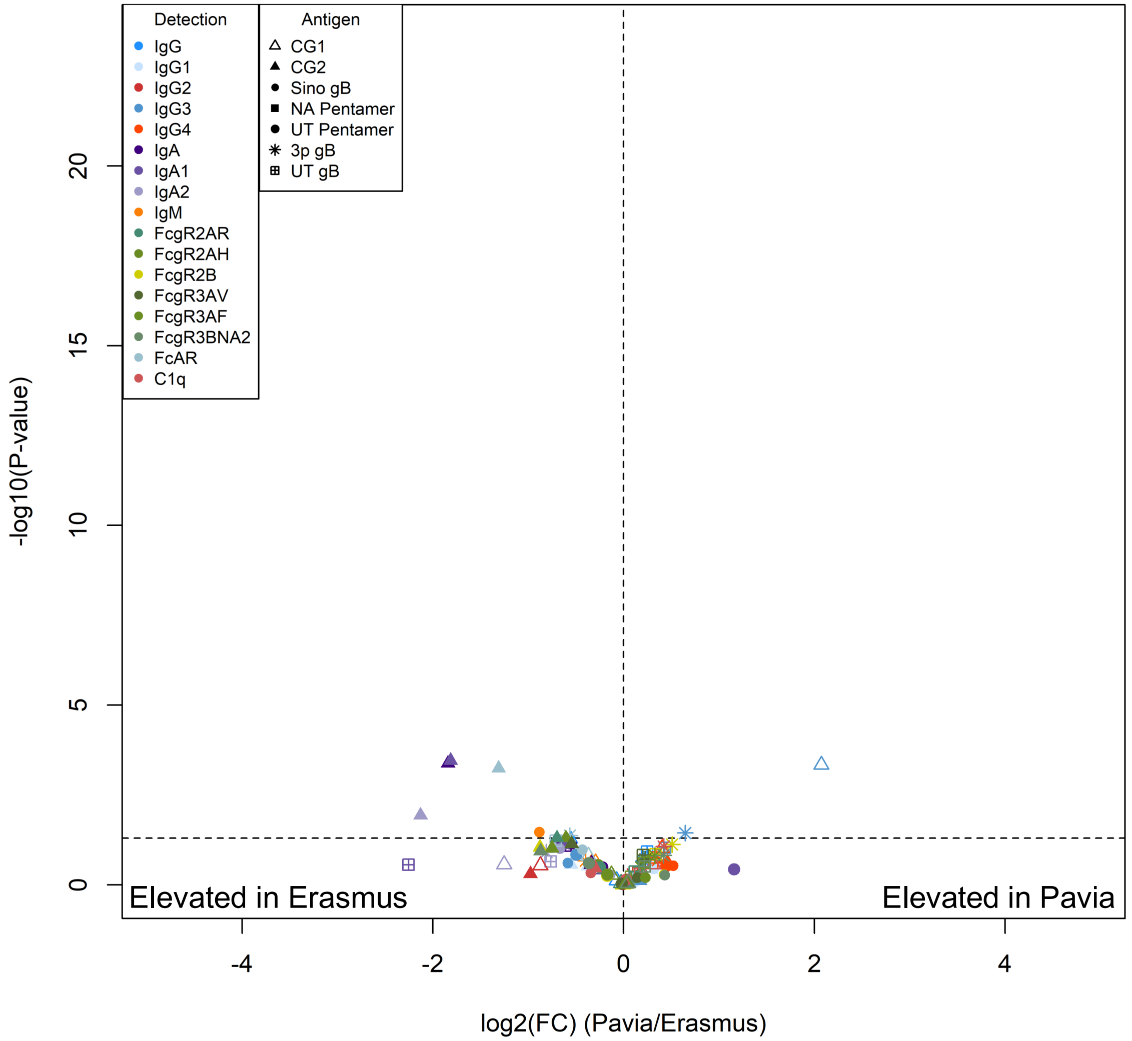


**Supplemental Figure 3. Comparison of antibody responses across medical centers.** Volcano plot of each CMV-specific antibody feature assessed. Volcano plot represents the log2 fold change (x-axis) against the –log10 p value (Mann Whitney test). Antibody specificities (Antigen) are indicated by shape and Fc characteristics (Detection) indicated by color.

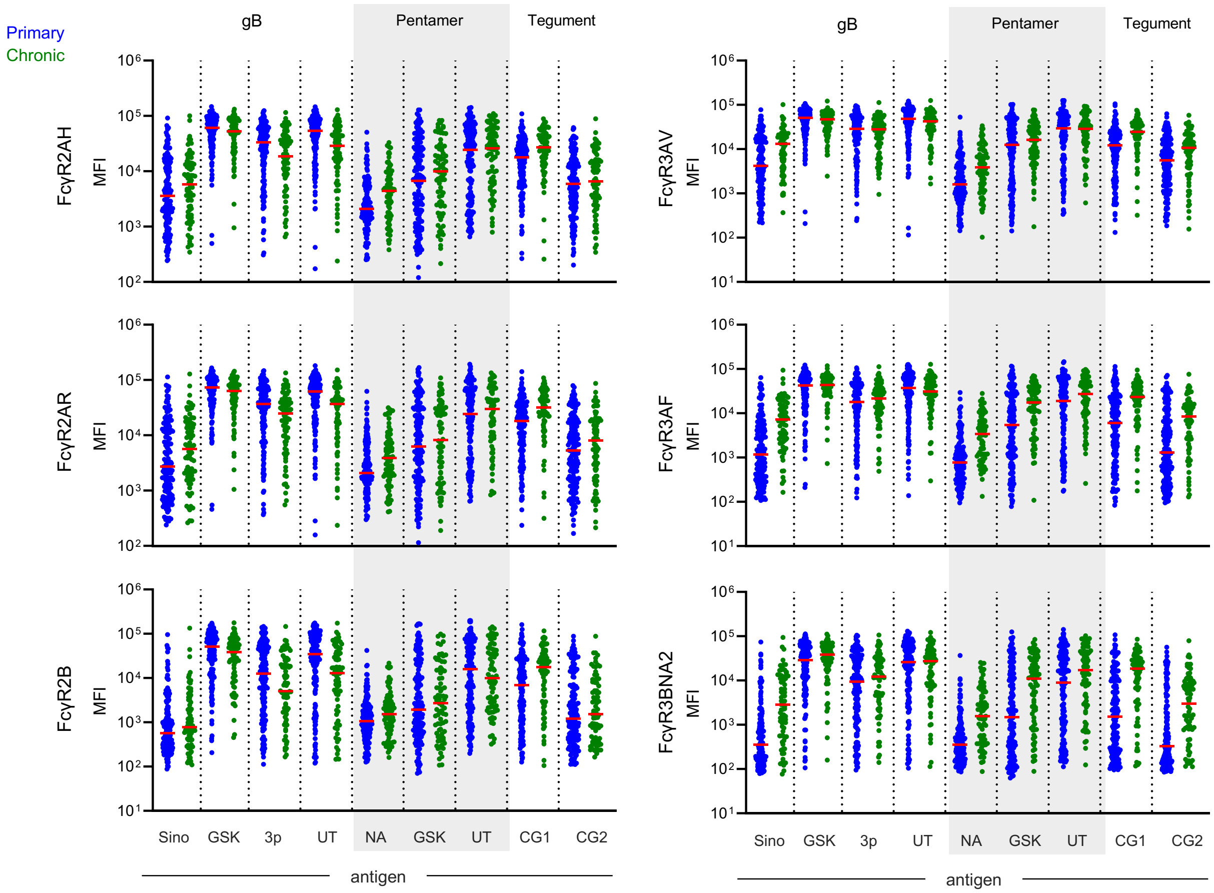


**Supplemental Figure 4. Representative boxplots of CMV-specific antibody binding to FcγR across groups.** FcγR2AH, FcγR2AR, FcγR2B, FcγR3AV, FcγR3AF, FcγR3BNA2 levels, as defined by median fluorescent intensity (MFI) in subjects with primary (blue) or chronic (green) CMV infection. Values represent the mean of technical replicates; bars indicate group medians.

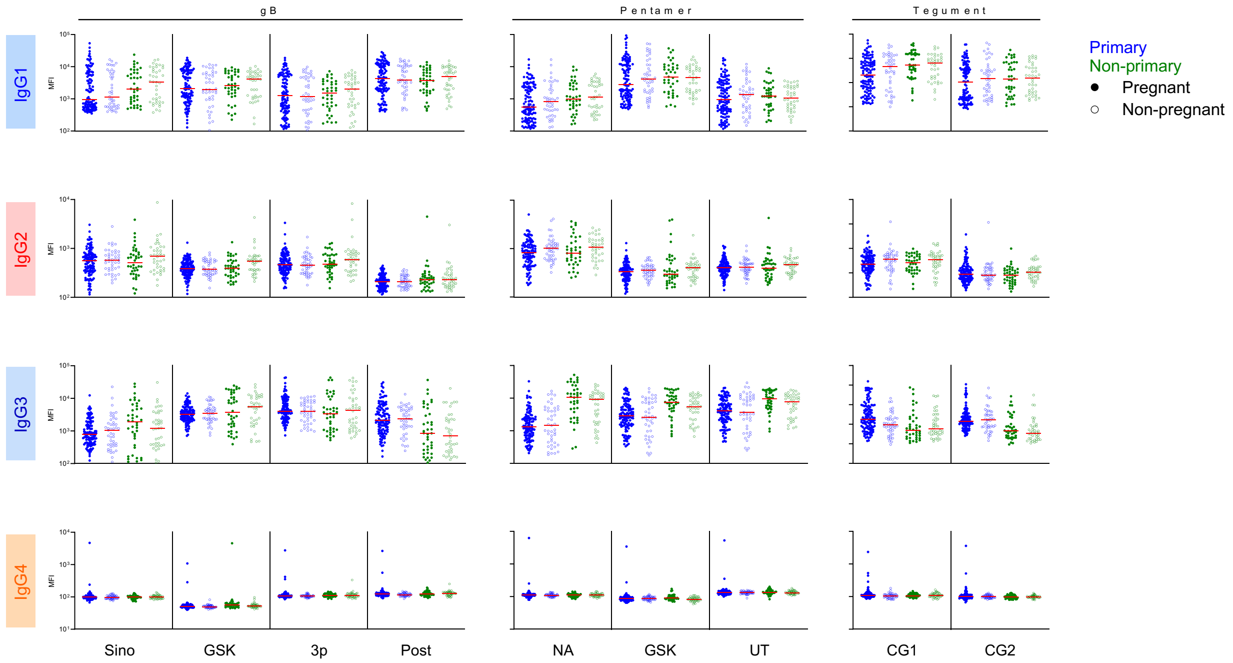


**Supplemental Figure 5. Representative boxplots of CMV-specific IgG subclasses across groups.** IgG1, IgG2, IgG3, and IgG4 levels, as defined by median fluorescent intensity (MFI) in pregnant (filled spheres) or non-pregnant (hollow spheres) subjects with primary (blue) or chronic (green) CMV infection. Values represent the mean of technical replicates; bars indicate group medians.

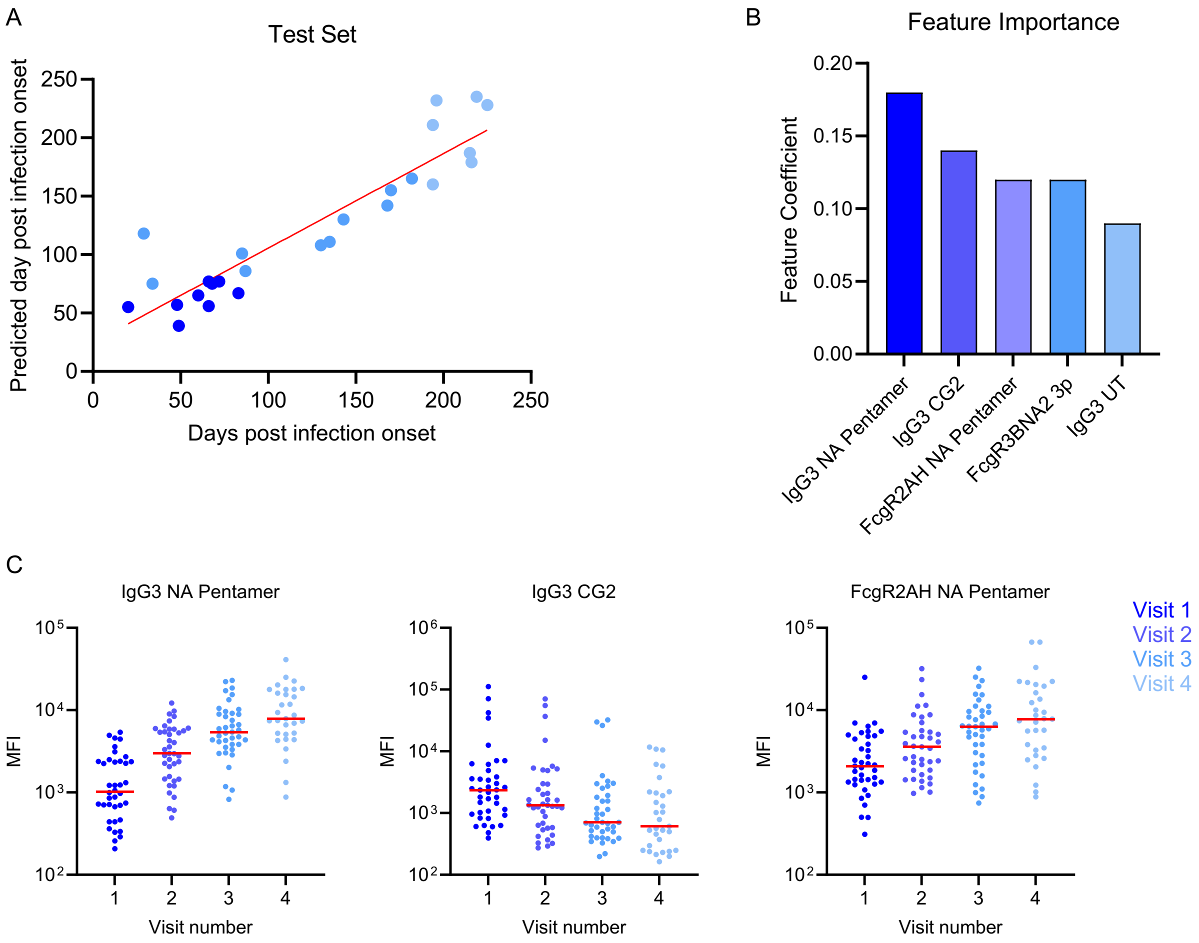


**Supplemental Figure 6. Longitudinal machine learning model performance and feature importance. A.** Representative test set for predicting days post infection. Symbols are colored according to visit number. Red line denotes the best fit line x=y. **B.** Top five model features according to feature importance. **C.** Boxplots of the top three model features plotted according to visit number.

**Supplemental Table 1**. Fc detection and antigen reagents

| **Antigen** | **Source** |
|---|---|
| **Other** | |
| Tetanus | Sigma 676570-37-9 |
| RSV dsCav1 | McLellan et al., Science. 2013 |
| Pertactin | VWR 102946-462 |
| Rubella capsid | Abcam ab43034 |
| Neuraminidase | Immune Technology IT-003-00110p |
| Hepatitis B | Zageno H1909-17C |
| **CMV** | |
| gB UT | Ye et al., PLoS Pathog. 2020 |
| gB 3p (JSM-956) | This study |
| gB GSK | Chandramouli et al., Nat Commun. 2015 |
| gB UT (JSM-1074) | Sino Biological 10202-V08H1 |
| Pentamer UT | Wrapp et al., Sci Adv. 2022 |
| Pentamer NA | Native Antigen CMV-PENT |
| Pentamer GSK | Chandramouli et al., Sci Immunol. 2017 |
| Tegument CG1 (pp150/2-pp52/3) | Vornhagen et al., J Clin Microbiol. 1994 |
| Tegument CG2 (pp150/7-pp150/1) | Vornhagen et al., J Clin Microbiol. 1994 |
| **Fc Detection** | **Source** |
| a- IgG | Southern Biotech 1030-09 |
| a-IgG1 | Southern Biotech 9054-09 |
| a-IgG2 | Southern Biotech 9070-09 |
| a-IgG3 | Southern Biotech 9210-09 |
| a-IgG4 | Southern Biotech 9200-09 |
| a-IgA | Southern Biotech 2050-09 |
| a-IgA1 | Southern Biotech 9130-09 |
| a-IgA2 | Southern Biotech 9140-09 |
| a-IgM | Southern Biotech 9020-09 |
| FcγRIIa R131 | Boesch et al., mAbs. 2014 |
| FcγRIIa H131 | Boesch et al., mAbs. 2014 |
| FcγRIIb | Boesch et al., mAbs. 2014 |
| FcγRIIIa V158 | Boesch et al., mAbs. 2014 |
| FcγRIIIa F158 | Boesch et al., mAbs. 2014 |
| FcγRIIIb NA2 | Boesch et al., mAbs. 2014 |
| FcαR | Duke Protein Production Facility |
| C1q | Sigma 204876 |
